# Gene-by-environment interactions involving maternal exposures with orofacial cleft risk in Filipinos

**DOI:** 10.1101/2024.12.16.24319123

**Authors:** Zeynep Erdogan-Yildirim, Jenna C. Carlson, Nandita Mukhopadhyay, Elizabeth J. Leslie, Carmencita Padilla, Jeffrey C. Murray, Terri H. Beaty, Seth M. Weinberg, Mary L. Marazita, John R. Shaffer

**Affiliations:** Center for Craniofacial and Dental Genetics, Department of Oral and Craniofacial Sciences, School of Dental Medicine, University of Pittsburgh, Pittsburgh, PA 15261, USA; Department of Human Genetics, School of Public Health, University of Pittsburgh, Pittsburgh, PA 15261, USA; Department of Biostatistics and Health Data Science, School of Public Health, University of Pittsburgh, Pittsburgh, PA 15261, USA; Department of Human Genetics, Emory University School of Medicine, Atlanta, GA 30322, USA; Department of Pediatrics, College of Medicine, Institute of Human Genetics, National Institutes of Health, University of the Philippines, Manila, the Philippines; Department of Pediatrics, University of Iowa School of Medicine, Iowa City, IA, 52242, USA; Department of Epidemiology, Bloomberg School of Public Health, John Hopkins University, Baltimore, MD 21205, USA; Departments of Psychiatry and Clinical and Translational Sciences, School of Medicine, University of Pittsburgh, Pittsburgh, PA 15261, USA

## Abstract

Maternal exposures are known to influence the risk of isolated cleft lip with or without cleft palate (CL/P) – a common and highly heritable birth defect with a multifactorial etiology. To identify new CL/P risk loci, we conducted a genome-wide gene-environment interaction (GEI) analysis of CL/P on a sample of 540 cases and 260 controls recruited from the Philippines, incorporating the interaction effects of genetic variants with maternal smoking and vitamin use. As GEI analyses are typically low in power and the results can be difficult to interpret, we used multiple testing frameworks to evaluate potential GEI effects: 1 degree-of-freedom (1df) GxE test, the 3df joint test, and the two-step EDGE approach. While we did not detect any genome-wide significant interactions, we detected 12 suggestive GEI with smoking and 25 suggestive GEI with vitamin use between all testing frameworks. Several of these loci showed biological plausibility. Notable interactions with smoking include loci near *FEZF1*, *TWIST2,* and *NET1.* While *FEZF1* is involved in early neuronal development, *TWIST2* and *NET1* regulate epithelial-mesenchymal transition which is required for proper lip and palate fusion. Interactions with vitamins encompass *CECR2*— a chromatin remodeling protein required for neural tube closure—and *FURIN,* a critical protease during early embryogenesis that activates various growth factor and extracellular-matrix protein. The activity of both proteins is influenced by folic acid. Our findings highlight the critical role of maternal exposures in identifying genes associated with structural birth defects such as CL/P and provide new paths to explore for CL/P genetics.

## Introduction

Orofacial clefts (OFCs) are common birth defects that result mainly from failure in the processes required for the complete fusion of the structures involving the lip and/or the palate. On average, 1 in 700 live births are affected by OFCs worldwide with significant variability in incidence rates across populations.^1,2^ In a majority of OFC cases, the condition presents as cleft lip with and without cleft palate (CL/P), with 70% manifesting as an isolated, non-syndromic feature^3^. The rate of isolated CL/P is high in the Philippines, with 1 in 500 newborns affected.^1^ Siblings of affected Filipinos, have an 11.5-fold increased CL/P risk compared to the general population, indicating a strong familial component.^1,2^ A significant disparity in CL/P risk is evident across socioeconomic status (SES), with incidence rate dropping to approximately 1 in 1000 among Filipinos living under higher SES, suggesting poor maternal nutrition could be a contributing factor.^1,4,5^

Epidemiological studies in individuals from the Philippines have identified links between increased CL/P risk and inadequate levels of vitamin B6 (< 20nmol/L) as well as low plasma zinc levels in mothers.^4–6^ Although maternal smoking is another risk factor for clefts, studies involving Filipino mothers did not show statistically significant associations between smoking and risk of cleft ^7^, which may be due to Southeast Asian women having the lowest prevalence of smoking (0.9%) compared to other continental groups (1.4% through 17.5%).^8^ However, household smoking, as a possible indicator of passive smoke exposure, increased the odds for OFCs in offsprings.^7,9^ These findings align with previous reports showing evidence for increased risk for OFC with exposure to maternal smoking, environmental tobacco smoke exposure, and with lack of multivitamin (with/without folic acid) supplementation during pregnancy.^10^ These facts underscore the significance of both the genetic predisposition and the influence of environmental factors in CL/P etiology in this population.^1^

Genetic studies of isolated CL/P in diverse populations, including both candidate and genome-wide approaches, have been used to identify genetic risk loci.^11,12^ Although numerous CL/P risk loci have been identified, they collectively do not fully account for the estimated genetic variance. Interactions between genetic and environmental factors during development may explain some of the missing heritability, hence, several studies have explored gene-environment interactions (GEIs) in cleft risk in different populations and detected significant interactions with maternal smoking (*NOS3,*^13^ *GRID2,*^14^ and *ELAVL2*^14^), environmental tobacco smoke (*RUNX2*^15^) and maternal vitamin use (*NOS3,*^13^ *CACNG3*,^16^ and *ESRRG*^17^). Several suggestive interactions have been reported with maternal smoking (*MUSK*^18^ and *PRL*^16^) and maternal vitamin intake (*RETREG1,*^18^ FLJ0838,^17^ *COBL*,^17^ *CASP9*,^18^ and *ANTXR1*^18^). These findings highlight the significance of GEI studies especially for complex disorders such as CL/P for unveiling underlying mechanisms that cannot be detected by examining the genetic main effects alone.

Relatively little is known about genetic factors underlying CL/P or their interactions with environmental exposures in Filipinos, despite being a population with a high CL/P prevalence and the documented prevalence differences by SES that are evidence of environmental effects. Hence, the goal of our study was to discover new genes involved in orofacial cleft risk through genome-wide interrogation of GEIs in individuals from the Philippines. In particular, we focus on maternal smoking and vitamin use during the periconceptional period.

## Materials and methods

### Cohort description

For this GEI analysis, we included a total of 800 individuals recruited from the Philippines, which we will refer to as the discovery sample (sample flowchart in Figure S1A). We selected CL/P cases and unaffected unrelated controls who had complete data on maternal smoking and vitamin intake during the periconceptional period that spans from 3 months prior to conception to the end of first trimester. An individual was considered exposed if the mother confirmed personal smoking or vitamin intake (1) within the three months prior to pregnancy and/or (2) within the first trimester. Cases with a family history of cleft palate or a diagnosis of a syndrome and controls reporting family history of any type of craniofacial anomaly were excluded from analysis. We also removed individuals with unknown maternal exposure status across both time periods. The final sample included 540 cases and 260 controls. Details on the exposure count are provided in Table 1.

**Table 1.** Descriptive characteristic of study participants in discovery and replication cohorts.

|  |  | Discovery |  | Replication 1 | Replication 2 | Replication 3 |
| --- | --- | --- | --- | --- | --- | --- |
|  |  | CL/P cases | Controls | CL/P cases | CL/P cases | CL/P cases |
| N |  | 540 | 260 | 137 | 88 | 213 |
| Maternal vitamin use | Yes | 204 | 222 | 108 | 20 | 36 |
|  | No | 303 | 38 | 29 | 63 | 170 |
|  | unknown | 33 | 0 | 0 | 5 | 7 |
| Maternal smoking | Yes | 72 | 23 | 10 | 4 | 33 |
|  | No | 468 | 237 | 124 | 84 | 180 |
|  | unknown | 0 | 0 | 3 | 0 | 0 |
| Assigned sex at birth | Males | 320 | 123 | 85 | 59 | 139 |
|  | Females | 220 | 137 | 52 | 29 | 74 |

The study protocol was approved by Institutional Review Boards locally (University of the Philippines, Manila, FWA00003505) and at the University of Pittsburgh (FWA00006790). Written informed consent was obtained from all individuals prior to study participation.

### Genotyping and imputation

Genotyping of the Filipino cohort used in this analysis was performed as part of the Pittsburgh Orofacial Cleft 2 (POFC2) study that included 4,114 participants recruited worldwide across South Asia, Africa, Latin America, USA, East Asia and Europe (Figure S1A). The genomic DNA was extracted from saliva samples collected with Oragene kits (DNA Genotek Inc., Canada) and genotyped at the Center for Inherited Disease Research (CIDR) at John Hopkins University using the Illumina Global Diversity Array-8 v1.0 (GDA) covering approximately 1.9 million markers. After extensive quality control and quality assurance steps^19^ (removing poorly performing samples and those with sex discrepancies and variants with missing call rates (≥ 2%), discordant calls on duplicate probes, ≥ 2 Mendelian errors, deviation from Hardy-Weinberg equilibrium (*p* < 3.45 × 10^-3^)). Imputation of unobserved variants was conducted via the TOPMed Imputation Server with the TOPMed reference panel (version r2) using minimac4 (v1.6.0).^20,21^ Monomorphic variants and variants with low imputation quality (*R*^2^ < 0.8) were removed prior to analysis. The imputed dosages were converted to binary dosage file format via the R package *BinaryDosage*.^22^

The genotype data are available via the database of Genotypes and Phenotypes (dbGaP) via the accession number phs002815.v2.p1; the phenotypic and pregnancy history data are available through FaceBase (facebase.org; Accession number: FB00001368; doi: 10.25550/56-ES6P and Accession number: FB00001369, doi: 10.25550/5A-FJBJ).

### Statistical analyses

#### Gene-by-environment interaction analyses

For discovering G×E interactions (GEI) with maternal smoking and vitamin intake implicated in CL/P risk, we performed a genome-wide scan on common variants with a minor allele frequency (MAF) ≥ 0.10 leveraging the analytical approaches implemented in the R package *GxEScanR.*^23,24^ We employed logistic regression on the imputed dosages adjusting for reported sex and the first five principal components of ancestry (Figure S2).

There are many existing statistical approaches to detect GEI and interpreting the results from statistical models for GEI can be challenging. Thus, we employed three complementary strategies as implemented in *GxEScanR* for detecting and interpreting GEI in this study (Figure 1). The first approach used the standard 1 degree-of-freedom (1df) G×E test from a model with both gene and environment main effects and their interaction. While 1df G×E test is the easiest to interpret, it has the lowest power. The second approach was a unified 3df joint test of the main genetic effect (DG) from a model without an interaction, the same G×E effect as the first approach, and the gene-environment (GE) association. Since 3df joint test combines these components, it does not distinguish which one is driving the association. Hence, we conclude the presence of GEI effect, if the 1df G×E test and/or if the GE association test in cases but not in controls has a *p* < 0.05. The 3df test is powerful to detect signals by any combination of the three components, however, when the true effect is driven only by one component, the power to detect is reduced. Lastly, our third approach, called the two-step EDGE approach, uses a 2df joint test of DG and GE to screen variants, followed by a test of for the same G×E effect as the first approach. This two-step EDGE approach ultimately tests the same effect as the first approach, but with more statistical power as the filtering step reduces the total number of tests in step 2. However, SNPs with weak marginal genetic or environmental effects may lead to missed true interactions.^23–25^

**Figure 1.**
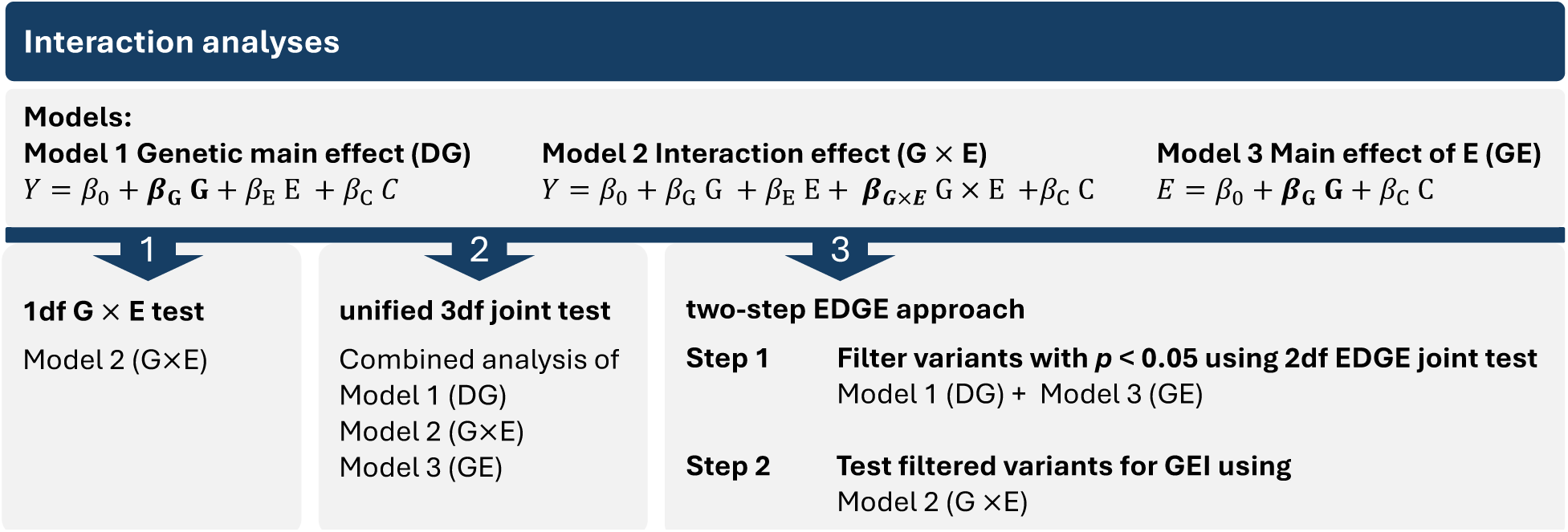
The three GEI analysis approaches used in this study. In the models, Y represents the phenotype, *β_G×E_* the multiplicative interaction effect, *β_G_* the genotypic effect, *β_E_* the environmental effect and C other covariates.

We used separate significance thresholds for each approach. For the first approach, our significance threshold was *p*_1df_ < 5 × 10^-8^. For the second approach (3df test), findings with a *p*_3df_ < 5 × 10^-8^/3 (β 2 × 10^-8^) were considered significant and associations with *p*_3df_ < 1 × 10^-5^ were considered suggestive. For our third approach (two-step EDGE), we used a filtering threshold of *p*_2df_EDGE_ < 0.05 for step 1, followed by a significance threshold of *p*_G×E_ β 8 × 10^-7^ for step 2 which was adjusted for the effective number of independent loci determined via simple.^23,26^ Step 2 associations with a *p*_G×E_ < 5 × 10^-4^ were considered suggestive.

### Regional plots

To investigate the genomic region of interest, we used LocusZoom^27^ to visualize the association signals within the context of nearby genes and regulatory elements. We incorporated data on strong craniofacial specific enhancers^28^ and the topologically associated domain information from H1-hESC Micro-C track available through UCSC Genome Browser into regional plots.^29^ To accurately reflect the genetic architecture of the study sample, we calculated the linkage disequilibrium in individuals from the Philippines in our discovery sample.

### Sensitivity analysis

Because maternal smoking and vitamin use are moderately correlated (Cramér’s V coefficient = 0.183, *p* = 3.81 × 10^-7^), to disentangle the effects of smoking, we reran the GEI analyses for vitamin use by excluding all individuals exposed to periconceptional smoking (N=99) leaving 676 individuals consisting of 439 CL/P cases (vitamin use: 44.19%) and 237 controls (vitamin use: 86.08%).

### Replication

To validate the GEI findings from the discovery cohort, we performed replication analysis with variants passing the suggestive threshold for the 3df test and/or the two-step approach. To reduce multiple testing burden at replication stage, we ran replication analysis for only those that demonstrated a GEI association from variants identified by the 3df test, either (1) via 1df G×E test with *p*_G×E_ < 0.05 or (2) via GE association in cases only with *p*_case_only_ < 0.05 with an insignificant GE association in controls.

We used a case-only replication approach using three independent sets of individuals with CL/P who were recruited from the Philippines (Table 1): *n* = 137 from the POFC1 Study conducted by the University of Pittsburgh (Replication 1)^30^, n = 88 from the GENEVA Study (Replication 2),^31^ and n = 213 from the Gabriella Miller Kids First (GMKF) initiative (Replication 3). Replication 2 and Replication 3 samples were originally enrolled in cleft studies conducted by the University of Iowa. Each of the three replication studies had differing family-based recruitment designs, so we extracted independent CL/P cases only for this replication analysis (removing overlapping and dependent individuals) so that the same statistical method could be used across each replication set (Figure S1B). Each study measured periconceptional exposure to maternal smoking and maternal vitamin use in the same way as the POFC2 cohort. In each replication sample separately, we evaluated GE association in cases to measure GEI with *GxEScanR*. We considered an interaction to be replicated if *p-*value < 0.05 in any replication sample with a consistent direction of effect between the discovery sample and the replication sample.

The genotyping for the POFC1 study (dbGaP accession number: phs000774.v2.p1) and the GENEVA study (dbGaP accession number: phs000094.v1.p1) were performed using Illumina HumanCore + Exome Array and Illumina Human 610 Quadv1_B array, respectively. More details about the genotyping for the POFC1 and the GENEVA studies has been described previously.^30,31^ We re-imputed both cohorts using the TOPMED r2 after removing low-frequency variants (MAF < 0.01) and phasing the haplotypes via SHAPEIT. The whole genome sequences from GMKF (dbGaP accession number: phs002595.v1.p1) were generated by the Broad Institute using Illumina HiSeqX with a target read depth of ∼30× coverage. The data were aligned to the GRCh38/hg38 reference genome and processed by the Kids First Data Research Center via Cavatica using a custom pipeline based on Genome Analysis Toolkit (GATK) best practices.^32^ The GATK genotyping workflow incorporated base quality score recalibration (BQSR), single-sample variant calling for SNVs and indels using HaplotypeCaller, joint variant calling across multiple samples, and final refinement with the variant quality score recalibration (VQSR) and filtering of called variants. Kids First DRC pipelines are publicly accessible as open-source tools on GitHub (Link for Alignment workflow and Joint genotyping workflow in Web Resources).

### Examination of previously known GEI loci implicated in CL/P risk in this GEI analysis

We explored our GEI analysis results for known significant statistically significant GEI loci with maternal smoking and vitamin intake reported in association with CL/P risk (Table 2). For each locus, we selected the lead SNV with the lowest p-value and considered it replicated if the *p*_G×E_ and/or *p*_case_only_ in our analysis was < 0.05.

**Table 2.** Look-up of previously known GEI loci with maternal smoking and vitamin intake implicated in CL/P risk in this GEI study.

| Exposure | Variant info |  |  |  |  |  | Discovery sample |  |  |  |  |  |  |
| --- | --- | --- | --- | --- | --- | --- | --- | --- | --- | --- | --- | --- | --- |
|  | SNV (hg38) | rsID | nearest gene | type | Study MAF | Reference | AF | GxE |  | DG |  | GE cases |  |
|  |  |  |  |  |  |  |  | OR | P | OR | P | OR | P |
| Smoking | 4-92455080-G-A | rs4389540 | GRID2 | intronic | 0.14 | Beaty et al (2013)<sup>14</sup> | 0.005 | - | - | - | - | - | - |
| Smoking | 7-150999023-T-G | rs1799983 | NOS3 | exonic - missense | 0.26 | Shaw et al (2005)<sup>13</sup> | 0.17 | 0.76 | 6.16E-01 | 0.99 | 9.33E-01 | 0.91 | 7.19E-01 |
| Smoking | 9-24527359-G-A | rs2257210 | ELAVL2 | intergenic | 0.31 | Beaty et al (2013)<sup>14</sup> | 0.21 | 1.11 | 8.23E-01 | 0.97 | 8.24E-01 | 1.31 | 2.30E-01 |
| Vitamin intake | 1-216999264-T-C | rs1339221 | ESRRG | intronic | 0.40 | Haaland et al (2018)<sup>17</sup> | 0.32 | 0.54 | 6.01E-02 | 1.03 | 8.33E-01 | 0.83 | 2.03E-01 |
| Vitamin intake | 7-150999023-T-G | rs1799983 | NOS3 | exonic - missense | 0.26 | Shaw et al (2005)<sup>13</sup> | 0.17 | 3.93 | 6.42E-03 | 0.85 | 3.60E-01 | 1.04 | 8.59E-01 |
| Vitamin intake | 16-24342036-A-G | rs9930171 | CACNG3 | intronic | 0.35 | Carlson et al (2022)<sup>16</sup> | 0.41 | 0.85 | 6.04E-01 | 0.93 | 5.92E-01 | 0.96 | 7.60E-01 |

## Results

In this study, we carried out a comprehensive analysis to detect GEIs with maternal smoking and vitamin use implicated in isolated CL/P risk in people recruited from Philippines. Maternal smoking showed a trend with increased risk of CL/P (OR = 1.58 [95%-CI = 0.95 – 2.73]), although the association was not statistically significant (*p* = 0.0796), and maternal vitamin intake was associated with reduced cleft risk (OR = 0.11[95%-CI = 0.08 – 0.17]; *p* < 2.2 × 10^-16^). The descriptive characteristics of the study sample are shown in Table 1.

### Genome-wide interaction analysis for maternal smoking

The statistical testing for GEI interaction with periconceptional exposure to smoking did not yield any genome-wide significant results using all three analytical approaches, however several suggestive associations were detected (Table S1). Our first statistical approach, the 1df G×E test, showed suggestive association with one variant (rs10150710-C, *p* = 5.90 × 10^-6^, Table S2) mapping to the second intron of *BCL11B* that encodes a transcription factor with key functions in maturation of T-cells, and neurological and craniofacial development.^33^ The second approach, the 3df test, detected 13 independent suggestive loci (*p* < 1 × 10^-5^) (Table S3 and Figure S3A). Four of these loci (*PTPRD*, *TTBK*, *SLFN12L,* and *TNS1*) showed moderate G×E interaction with a *p*_G×E_ < 0.05 (*p*-values ranging from 7.5 × 10^-4^ to 2.2 × 10^-2^) and one locus, *FEZF1*, indicated interaction via the case-only analysis (*p*_case_only_ = 4.87 × 10^-4^, Table 3).

**Table 3.**
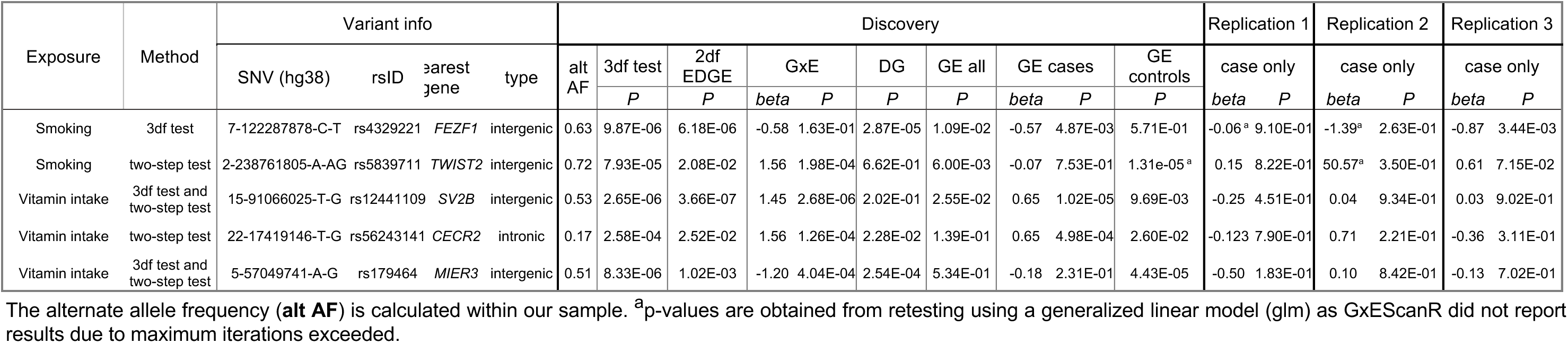
Main GEI findings with exposures to maternal smoking and maternal vitamin intake.

| Exposure | Method | Variant info |  |  |  | Discovery |  |  |  |  |  |  |  |  |  | Replication 1 |  | Replication 2 |  | Replication 3 |  |
| --- | --- | --- | --- | --- | --- | --- | --- | --- | --- | --- | --- | --- | --- | --- | --- | --- | --- | --- | --- | --- | --- |
|  |  | SNV (hg38) | rsID | nearest gene | type | alt AF | 3df test | 2df EDGE | GxE |  | DG | GE all | GE cases |  | GE controls | case only |  | case only |  | case only |  |
|  |  |  |  |  |  |  | P | P | beta | P | P | P | beta | P | P | beta | P | beta | P | beta | P |
| Smoking | 3df test | 7-122287878-C-T | rs4329221 | FEZF1 | intergenic | 0.63 | 9.87E-06 | 6.18E-06 | -0.58 | 1.63E-01 | 2.87E-05 | 1.09E-02 | -0.57 | 4.87E-03 | 5.71E-01 | -0.06<sup>a</sup> | 9.10E-01 | -1.39<sup>a</sup> | 2.63E-01 | -0.87 | 3.44E-03 |
| Smoking | two-step test | 2-238761805-A-AG | rs5839711 | TWIST2 | intergenic | 0.72 | 7.93E-05 | 2.08E-02 | 1.56 | 1.98E-04 | 6.62E-01 | 6.00E-03 | -0.07 | 7.53E-01 | 1.31E-05<sup>a</sup> | 0.15 | 8.22E-01 | 50.57<sup>a</sup> | 3.50E-01 | 0.61 | 7.15E-02 |
| Vitamin intake | 3df test and two-step test | 15-91066025-T-G | rs12441109 | SV2B | intergenic | 0.53 | 2.65E-06 | 3.66E-07 | 1.45 | 2.68E-06 | 2.02E-01 | 2.55E-02 | 0.65 | 1.02E-05 | 9.69E-03 | -0.25 | 4.51E-01 | 0.04 | 9.34E-01 | 0.03 | 9.02E-01 |
| Vitamin intake | two-step test | 22-17419146-T-G | rs56243141 | CECR2 | intronic | 0.17 | 2.58E-04 | 2.52E-02 | 1.56 | 1.26E-04 | 2.28E-02 | 1.39E-01 | 0.65 | 4.98E-04 | 2.60E-02 | -0.123 | 7.90E-01 | 0.71 | 2.21E-01 | -0.36 | 3.11E-01 |
| Vitamin intake | 3df test and two-step test | 5-57049741-A-G | rs179464 | MIER3 | intergenic | 0.51 | 8.33E-06 | 1.02E-03 | -1.20 | 4.04E-04 | 2.54E-04 | 5.34E-01 | -0.18 | 2.31E-01 | 4.43E-05 | -0.50 | 1.83E-01 | 0.10 | 8.42E-01 | -0.13 | 7.02E-01 |

The alternate allele frequency (**alt AF**) is calculated within our sample. <sup>a</sup>p-values are obtained from retesting using a generalized linear model (glm) as GxEScanR did not report results due to maximum iterations exceeded.

Using the two-step EDGE approach, we found another five suggestively associated risk loci (*PAMR1, SPAG16, TWIST2, NET1,* and *ZNF722,* Table S4 and Figure S3B). Two of these loci, *TWIST2* and *NET1* are implicated in the regulation of epithelial-mesenchymal transition (EMT), a critical process in lip and palate fusion during embryonic development. The lead signal at *TWIST2* (rs5839711-AG, *p_G×E_*= 2.0 × 10^-4^) is an intergenic variant (Figure 2B). This variant does not show evidence of a genetic main effect overall or when restricting to just the unexposed CL/P cases; however, among those with exposure to maternal smoking, the CL/P risk is higher in individuals carrying one or two copies of the inserted G allele compared to homozygotes for the A allele (Figure 3B). Per Genotype-Tissue Expression (GTEx^34^ v8) project data, this variant regulates the alternative splicing of the nearby lincRNA *AC144525.1* in transformed fibroblast cells (*p* = 1.55 × 10^-25^), where each copy of the alternate allele is associated with decreasing intron-excision ratio (Figure 4B). Additionally, this lincRNA is located within the enhancer region GH02J238784 according to the GeneHancer,^35^ that also encompasses the craniofacial-specific enhancers^28^ as shown in Figure 2B and exhibits regulatory effects on the surrounding genes including *TWIST2*, *LINC01937* and *ASB1*.

**Figure 2.**
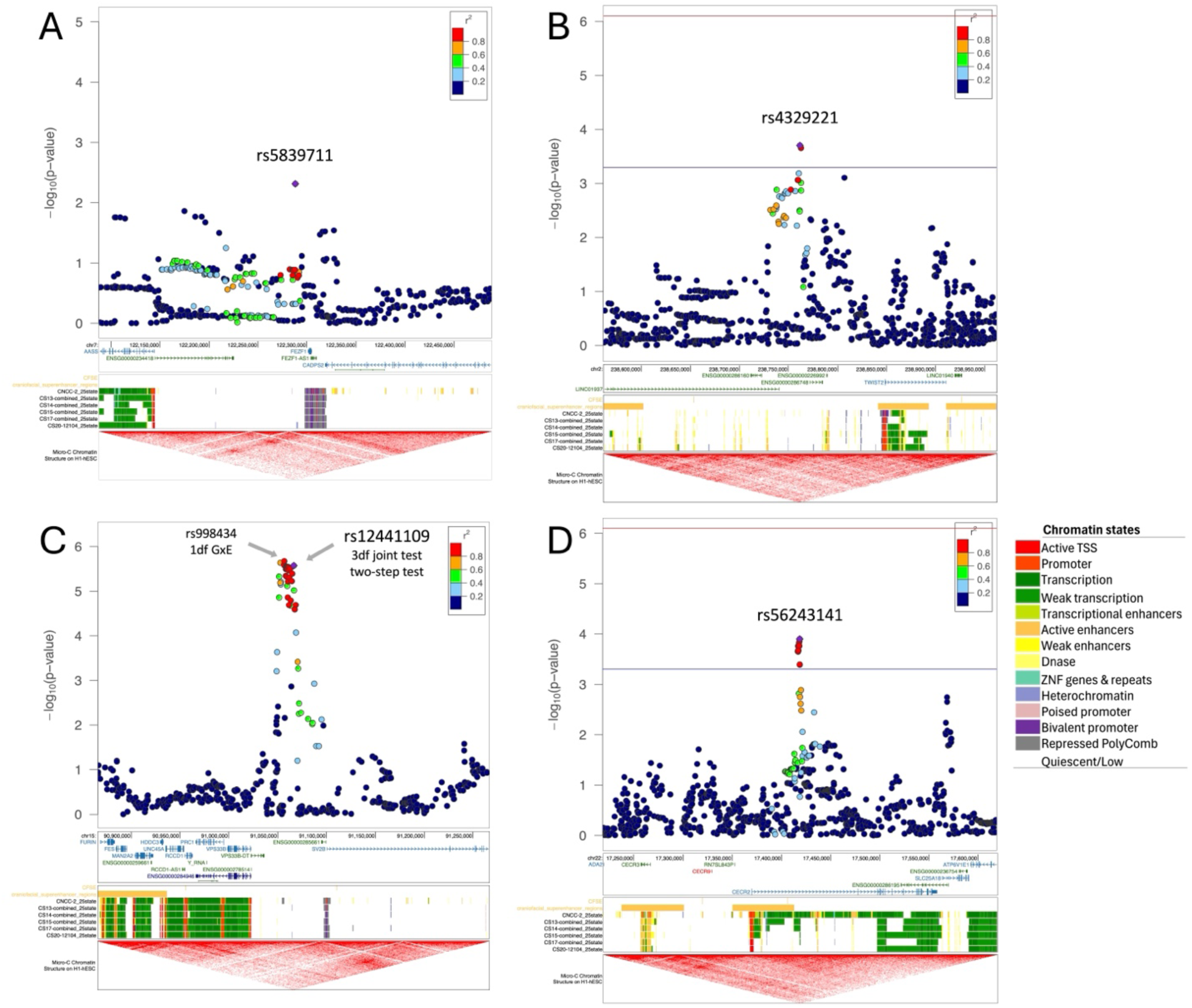
Regional plots for the main findings with maternal smoking (top) and maternal vitamin intake (bottom). The regional plots depict the genomic location of the identified variants (lead variant as purple diamond) in respect to nearby genes, the craniofacial enhancers and the topologically associated domains from UCSC genome browser track. For **(A)** rs5839711 at *FEZF1* locus, which was identified via 3df test, we display the –log_10_ of the p-values from case-only analysis. For **(B)** rs4329221-T, **(C)** rs12441109-G and **(D)** rs56243141 we report the –log_10_ p-values from 1df G×E test. For (B) and (D), which were identified via two-step testing, we provide the significance (red) and suggestive (blue) threshold for the second step. The regional plot for the locus at SV2B (C) depicts in addition to the main finding rs12441109-G via 3df test and two-step test, the lead variant rs998434-T from 1df G×E test. The colors of the variants indicate their correlation (r^2^) with the lead variant.

**Figure 3.**
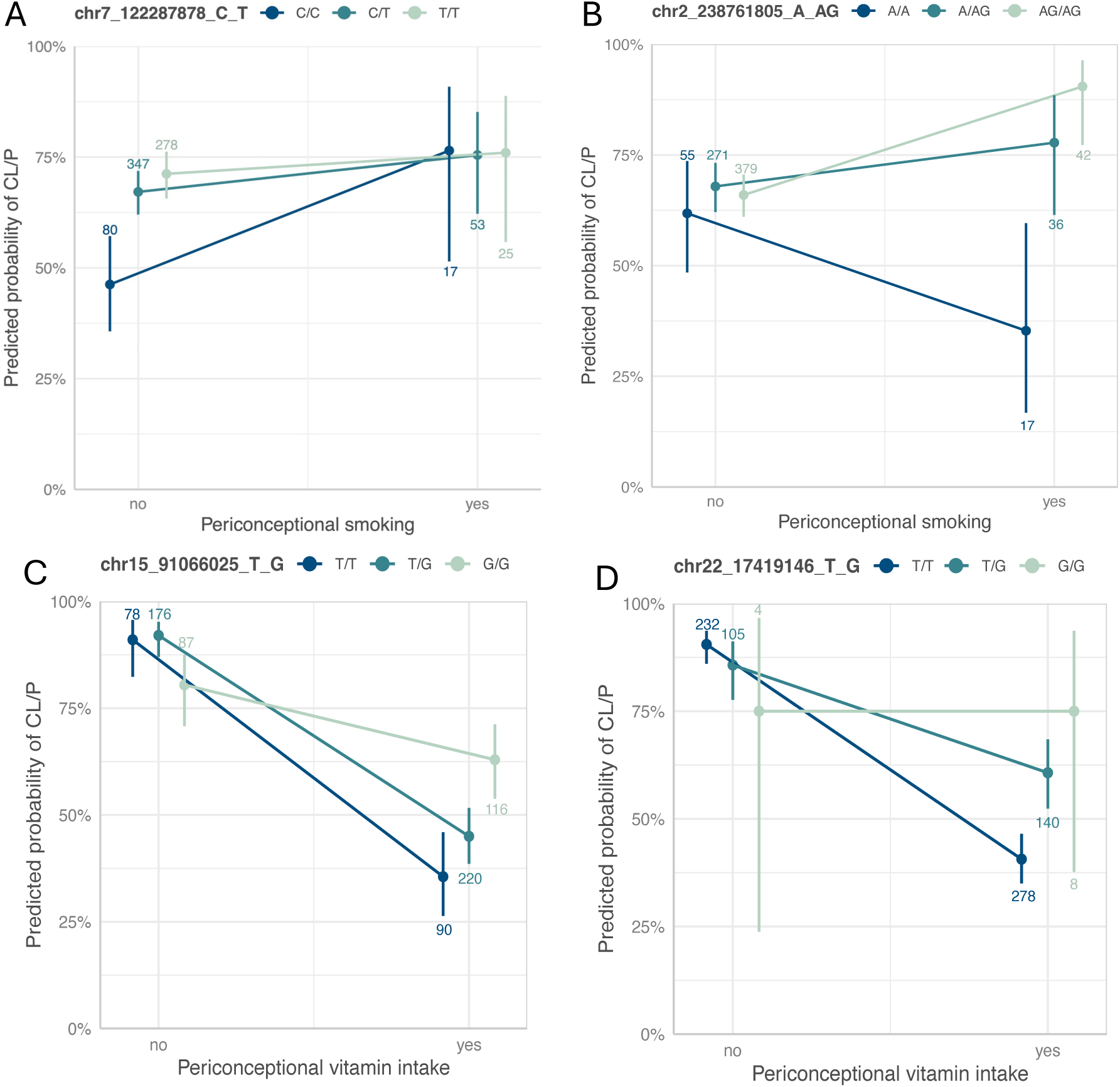
Interaction plots for the main findings with maternal smoking (top) and maternal vitamin use (bottom). Interaction plots between **(A)** rs4329221-T in *FEZF1* locus, and **(B)** rs5839711-AG in *TWIST2* locus and maternal smoking, and between **(C)** rs12441109-G in *SV2B* locus, and **(D)** rs56243141-G in *CECR2* locus with maternal vitamin use. Each panel is annotated with the chromosomal positional in hg38 followed by the reference and the alternate allele. The color scheme ranges from dark (homozygous for the reference allele) to light (homozygous for the alternate allele). The count of individuals within each stratum and genotype is annotated.

**Figure 4.**
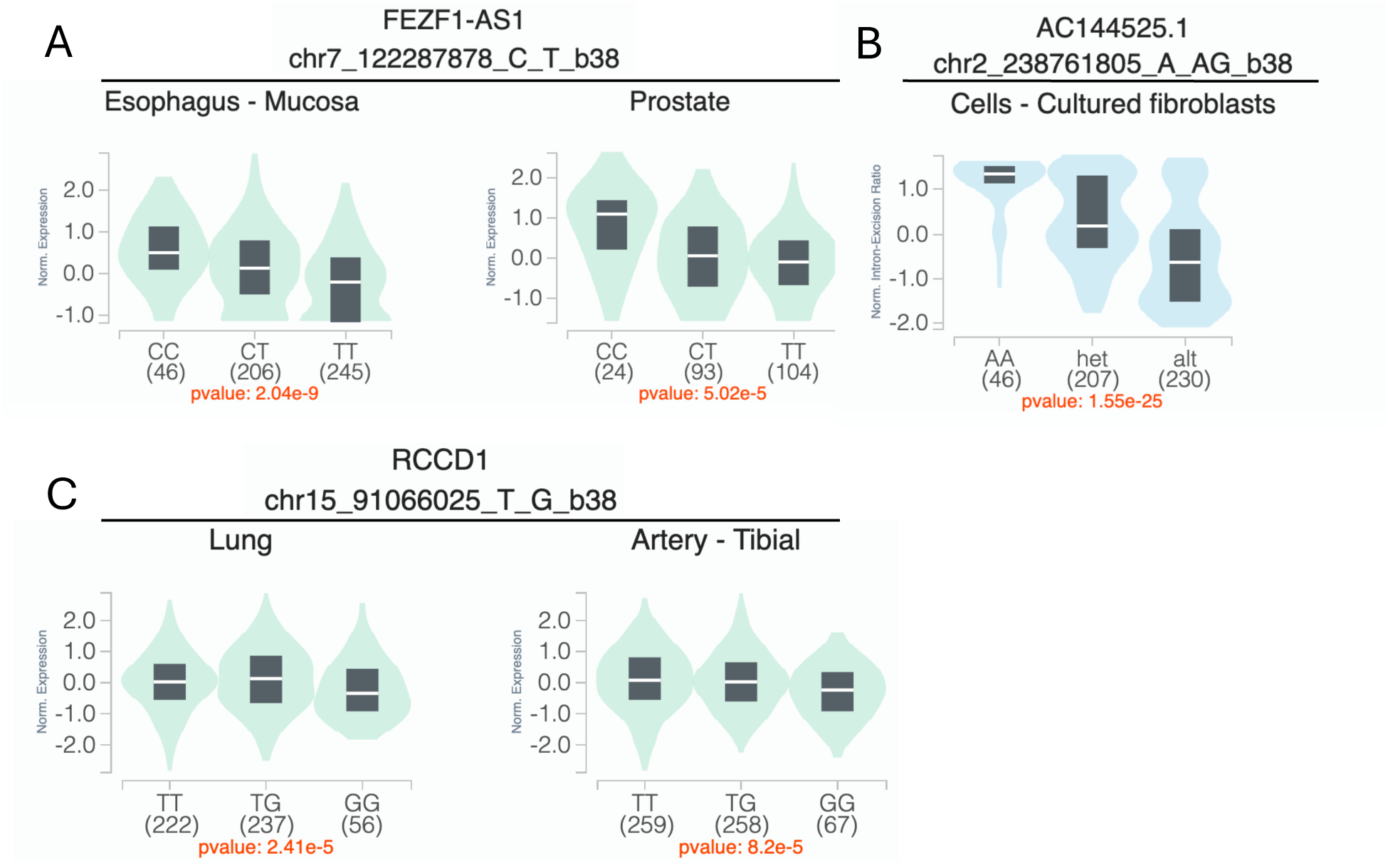
Variant-specific expression (eQTL) and splicing quantitative trait loci (sQTL) for the main findings with maternal smoking (top) and maternal vitamin use (bottom). (A) rs4329221-T is associated with reduced mean expression of *FEZF1-AS1* in both the esophageal mucosa (β = -0.36, *p* = 2.0 × 10^-9^) and the prostate tissue (β = -0.38, *p* = 5.0 × 10^-5^). (B) rs5839711-AG significantly decreases the intron-excision ratio of the lincRNA *AC144525.1,* which acts as enhancer on the nearby *TWIST2, ASB1* and *LINC01937,* in the cultured fibroblast cells (β = -0.78, *p* = 1.5 × 10^-25^). (**C**) rs12441109-G in *SV2B* locus reduces the mean expression of *RCCD1* in lung cells (β = -0.12, *p* = 2.4 × 10^-5^) and tibial artery (β = -0.11, *p* = 8.2 × 10^-5^). eQTL and sQTL data were retrieved from the GTEx browser.

### Genome-wide interaction analysis for maternal vitamin use

For the vitamin GEI analysis, while no variant achieved statistical significance using any of the three approaches, we identified four suggestive loci (*NLGN1, SV2B, CSMD1,* and *MIER3*) with moderate G×E effect via 3df test (Figure S4A) and 22 suggestive loci via two-step approach (Table S1, Figure S4B).

Using the 1df G×E test, the most significant association was at 15q26.1 in the intergenic region proximal to *SV2B*, with rs998434 (*p*_GxE_ = 2.13 × 10^-6^) as the top signal (Figure 2C, Table S5). The same locus was identified by both the 3df test and the two-step approach (Table 3, Table S6 and S7), through association with a variant in high LD (rs12441109-G, *p*_3df_ = 2.65 × 10^-6^ and *p*_GxE_ = 2.68 × 10^-6^, LD with rs998434 r^2^ = 0.95). While we did not observe a genetic main effect, rs12441109-G exhibited suggestive interaction with vitamin exposure. The protective effect of maternal vitamin use on CL/P risk differed by genotype with an estimated probability of CL/P of ∼58% for GG individuals with maternal vitamin use compared to ∼33% for TT (Figure 3C). For rs12441109-G, we observed a lower p-value after performing a sensitivity analysis by excluding smokers (*p*_3df_ = 9.05 × 10^-7^ and *p*_GxE_ = 9.34 × 10^-7^, Table S8). Furthermore, rs12441109-G acts as an eQTL on *RCCD1,* where the double copy of the G allele reduces the mean *RCCD1* expression in the tibial artery (*p* = 8.2 × 10^-5^) and the lung tissue (*p* = 2.41 × 10^-5^) per GTEx (Figure 4C).

Among the 22 loci detected via the two-stage approach (Figure S4B), one of the noteworthy findings was the *CECR2* locus with the lead variant rs56243141-G (*p*_GxE_ = 1.26 × 10^-4^, sensitivity analysis *p*_GxE_ = 1.34 × 10^-3^) mapping to its first intron (Table 3 and Table S7). When stratified by vitamin use, unexposed individuals with the TT genotype had higher estimated odds of CL/P although not statistically significant, whereas exposed individuals with the TT genotype had significantly lower odds of CL/P compared to the genotypes with the alternate allele G (Figure 3D). The odds of CL/P for individuals homozygous for the minor allele (G) remained same across both strata, however with wide confidence intervals as there were few homozygotes.

### Replication analysis

To validate our findings, we conducted a replication analysis across three independent samples consisting of individuals from the Philippines (Replication 1, Replication 2, and Replication 3). From the 39 suggestive findings, only rs4329221-T at the intergenic region near *FEZF1* replicated (Table 2 and Table S1). This variant demonstrated GEI effect with maternal smoking and was detected via the 3df test (effect estimate: -0.57, *p_case_only_* = 4.87 × 10^-3^). The interaction between rs4329221-T and smoking replicated in the Replication 3 sample with an effect estimate of -0.87 and *p_case_only_* = 3.44 × 10^-3^. Although this variant did not replicate in Replication 1 and Replication 2 samples, it demonstrated same direction of effect. While the CL/P risk in smokers was almost identical across all genotype strata, in non-smokers the two copies of the reference allele (CC) was associated with lower odds of CL/P compared to T carriers (Figure 3A). This variant also serves as an eQTL and leads to reduced expression of *FEZF1-AS1* in the esophageal mucosa and the prostate (*p* = 2.04 × 10^-9^ and *p* = 5.02 × 10^-5^, respectively) in carriers of the T allele (Figure 4A).

### Known GEI loci with maternal smoking and vitamin intake in CL/P risk

From six of the previously known significant GEIs, we replicated the interaction between *NOS3* and maternal vitamin intake. The candidate study by Shaw et al (2005) investigating the interaction between *NOS3* loci and maternal smoking and/or vitamin intake detected increased CL/P risk with rs1799983-T in offsprings who were exposed to maternal smoking without maternal vitamin supplementation during periconceptional period (OR = 4.4, 95%-CI [1.8, 10.7]).^13^ In our study, offsprings presenting with same exposure and genotypes were all affected (Table 2). When stratified by vitamin in the complete sample or in the subset excluding smokers, the predicted probability of CL/P was highest in non-exposed offsprings carrying the T allele and lowest in exposed individuals (1.00, 95%-CI [0, 1] vs 0.30, 95%-CI [0.10, 0.65]]. In contrast, although offsprings homozygous for the G allele demonstrated an advantage when their mothers did not take vitamins, there was no statistically significant difference in predicted probability among offsprings whose mothers used vitamins (G/G 0.49 [0.43, 0.55] and T/G 0.47 [0.39, 0.55]).

## Discussion

The goal of this study was to identify genetic risk variants that interact with well-established environmental risk factors, specifically periconceptional exposure to smoking and vitamin use, contributing to isolated CL/P in individuals from Philippines. To achieve this, we performed a comprehensive analysis of GEIs using a genome-wide approach by applying three complementary methods: the 1df G×E test, the unified 3df test and the two-step EDGE approach. While neither method identified genome-wide significant associations in our sample, our findings revealed 10 suggestive loci associated with maternal smoking and 25 loci associated with maternal vitamin use during periconceptional period. Among these suggestive loci, the GEI effect at the *FEZF1* locus was replicated in the three independent samples analyzed (Table 3). We also noted several of the remaining GEI findings for potential biological relevance to cleft risk: *TWIST2, NET1, TNS1*, and *BCL11B* for maternal smoking and *SV2B, CECR2,* and *MIER3* for maternal vitamin use. None of these findings has been not reported in previous GEI analyses of CL/P.^14,16–18,36–39^

Our main finding was that the *FEZF1* locus influenced CL/P risk via its interaction with smoking. *FEZF1* encodes a zinc finger protein that serves as a transcriptional repressor.^40^ *FEZF1* is an important factor for neuronal differentiation and migration of olfactory sensory neurons.^40^ The disruption in the maturation process of olfactory neurons, which is intertwined with the migration of gonadotropin releasing hormone (GnRH) neurons, are associated with hypogonadotropic hypogonadism 22 with anosmia (also known as Kallmann syndrome) or without anosmia (HH22 [MIM: 616030]) which have been reported with missense mutations within *FEZF1*.^40–42^ The lead variant rs4329221-T is an eQTL for the nearby located *FEZF1-AS1* (Figure 4).^34^ Its expression is reduced in carriers of the alternate allele (T).^34^ Furthermore, *FEZF1-AS1*, which promotes cell proliferation and inhibits apoptosis, is found overexpressed in placental tissues collected from preeclampsia patients and in various tumors predicting poor outcome.^43,44^ Moreover, the lead variant is 30kb proximal to the variants associated with smoking initiation (rs1443753-C^45^ and rs10953957-A^45^) and ever vs. never smokers (rs1443753-C,^45^ rs10252114-C,^46^ and rs10953957-A^47^).

Among the GEI loci identified in association with smoking, *TWIST2* is another potential candidate with biological role in CL/P risk. *TWIST2* encodes a basic helix-loop-helix type transcription factor that modulates the chromatin binding activity and has bifunctional role both as a transcriptional repressor or activator.^48^ *TWIST2* governs the mesenchymal cell fate and epithelial-mesenchymal transition (EMT) critical for normal embryonic morphogenesis and cancer progression.^49^ It is expressed in the mesodermal tissues (including craniofacial mesenchyme, osteoblasts, myocytes and adipocytes) during embryogenesis and prevents them from reaching their terminal differentiation.^48,50^ Mutations that disrupt these processes are found to cause facial patterning defects as seen in ablepharon macrostomia and Barber-Say syndrome.^49,51^ Additionally, *TWIST2* overexpression in breast cancers lead to downregulation of *CDH1* by repressing its promoter.^52^ Loss of *CDH1*/E-cadherin in neural crest cells impairs their migration leading to craniofacial malformations including CL/P in animal models that recapitulated the observations in CL/P patients with rare *CDH1* mutations.^53–56^ Furthermore, experiments with continuous exposure to smoke extract over 21 and 40 weeks increased the expression of *TWIST2* and reduced the expression of *CDH1* in mammary epithelial cells (non-tumorigenic MCF10A and tumorigenic MCF-7) promoting the migratory ability of the cells.^57^ Given its involvement in key developmental processes, *TWIST2* is a potential candidate for further investigation to determine its role with smoking in CL/P.

Analysis with another key exposure, the maternal vitamin intake during periconception, revealed several genes that stand out for their potential relevance to CL/P, such as *SV2B*. Multiple variants with GEI effect span the intergenic region between *SV2B* and *VPS33B* (Figure 2C), which is distally located to a large region of craniofacial enhancers that are linked by the same topologically associated domain (TAD). The lead variant rs12441109-G exhibits regulatory role on the expression of the neighboring gene *RCCD1* (Figure 4C), that is involved in chromatin organization and is recognized as a novel oncogene in lung cancer.^34,58^ RCCD1 promotes the migration of tumor cells and TGF-beta-induced EMT.^58^ Within the same TAD, *FURIN* is another potential candidate gene underlying the observed GEI effect. *FURIN* encodes a critical serine protease that facilitates the conversion of wide range of protein precursors, including growth factors, their receptors and extracellular matrix proteins, into active forms and plays vital role in early embryonic development. This is highlighted by the evidence that mouse embryos lacking *Furin* do not survive beyond 10-11 days after birth and present with notable defects in ventral closure and axial rotation which are under the control of BMP subfamily, to which the FURIN substrates TGFβ and related proteins including BMP4, and Nodal precursors belong. Moreover, FURIN activity is shown to be inhibited by folic acid.^59^ In addition to its mapping onto craniofacial-specific enhancers, another piece of evidence supporting its role for CL/P risk is that mutations in *GDF11*, which prevents its cleavage by FURIN, lead to CL/P.^60,61^

A second promising GEI locus with vitamin use is the *CECR2* that is responsible for chromatin remodeling and proper development of the neural tube and craniofacial structures. It is also reported in association with cat eye syndrome (CES [MIM: 115470]) that is caused by an inverted duplication involving the region at *CECR2*.^62^ Loss of *CECR2* in mice was shown to cause neural tube defects.^63^ Supplementation of folic acid in women during periconceptional period is a well-established preventative measure against neural tube defects. Similarly, maternal folic acid use also reduces occurrence as well as recurrence risk of OFCs.^64^ Studies have shown that folate increased the *CECR2* expression, while folate deficiency and the resulting increase in homocysteine levels, especially its metabolite homocysteine thiolactone, led to decreased expression of *CECR2*.^65^ Mutations within *CECR2* have been reported to curtail its expression.^63,65^ While there is evidence for interaction between the *CECR2* and folic acid, the mechanistic link to CL/P risk is unknown.

We also reviewed the literature for known GEI loci with maternal smoking and vitamin intake and limited the look up in our analysis only to the prior significant findings. We validated the association between the *NOS3* variant rs1799983-G and maternal vitamin intake in our discovery sample. Although we confirmed one of the GEI findings in our sample, replicating GEI findings remains challenging due to heterogeneity across study samples, populations, and how exposure status is assessed.

### Strengths

One of the major strengths of our study is the use of complementary GEI detection methods (1df GxE test, the 3df test and the two-step EDGE approach) that allowed us a robust assessment of potential interactions that might have been not captured by a single method. Furthermore, this study uniquely contributes to the genetic research by focusing on Filipino population, which is among the populations with highest CL/P risk and has a high level of exposure to adverse environmental factors as reported.^1^ Moreover, the examination of the interaction with vitamin intake (with/without folic acid) specifically within this population provides insights into the interaction as there was no fortification of food with folic acid at the time of recruitment.^66^

### Limitations

A major limitation of our study lies in the sample size, as GEI analyses generally require larger sample sizes than studies that examine the main genetic effects. Given the relatively small sample size of our sample, the study may have been underpowered to identify significant interactions, particularly for exposures with low prevalence such as smoking in Filipino women. Another limitation of our study may lie in the potential recall and reporting bias associated with exposure data, as these were collected retrospectively. There was also substantial heterogeneity in how exposure data are defined and collected and rates of exposures (especially vitamin use) across different study samples.

### Summary

In summary, we conducted a comprehensive genome-wide gene-by-environment analysis with maternal smoking and vitamin intake influencing CL/P risk in individuals from Philippines and identified several GEI loci that had not been reported before. These include notable interactions with smoking near *FEZF1* and *TWIST2*, and with vitamin intake within *CECR2* and near *FURIN*, all of which demonstrate biological plausibility and require further investigation.

## Description of supplemental information

Supplemental information includes one PDF file with four figures and an excel file with nine tables.

## Acknowledgement

The authors thank the study participants for contributing to this study. The authors are also grateful for the support from the National Institute of Health (NIH) funded through: R01-DE032122 (PI: Shaffer JR), X01-HG011437 (PI: Shaffer JR, Marazita ML), R01-DE016148 (PI: Marazita ML, Weinberg SM), X01-HG000784 (PI: Marazita ML), R37-DE008559 (PI: Murray JC), U01-DE018993 (PI: Beaty TH), and X01-HD100701-01(PI: Leslie EJ, Marazita ML, Murray, JC). Research reported in this publication was supported by the National Institute of Dental & Craniofacial Research of the National Institutes of Health under Award Number T90-DE030853 (PI: Sfeir C). The content is solely the responsibility of the authors and does not necessarily represent the official views of the National Institutes of Health. This research was also supported in part by the University of Pittsburgh Center for Research Computing (CRC), RRID: SCR_022735, through the resources provided. Specifically, this work used the HTC cluster, which is supported by NIH award number S10OD028483.

## Author contributions

**Conceptualization**, Z.E.-Y., J.C.C., J.R.S.; **Methodology**, Z.E.-Y., J.C.C., **Formal Analysis**, Z.E.-Y.; **Visualization**, Z.E.-Y.; **Writing** – **Original Draft**, Z.E.-Y.; **Writing – Review and Editing**, Z.E.-Y., J.R.S. S.M.W., J.C.C., M.L.M., E.J.L, N.M.; **Resources,** E.J.L., C.P., J.C.M., T.H.B., S.M.W., M.L.M., J.R.S.; **Supervision**, S.M.W., M.L.M., J.R.S.; **Funding Acquisition**, E.J.L., C.P., J.C.M., T.H.B., S.M.W., M.L.M., J.R.S.;

## Web resources

GTEx: https://gtexportal.org/

GxEScanR: https://github.com/USCbiostats/GxEScanR

Kids First DRC Alignment workflow: https://github.com/kids-first/kf-alignment-workflow

Kids First DRC Joint genotyping workflow: https://github.com/kids-first/kf-jointgenotyping-workflow

OMIM: http://www.omim.org

UCSC Browser: https://genome.ucsc.edu

## Data availability

All phenotype and genotype data used in this study are accessible via FaceBase and dbGaP.

**POFC2 Study:** The demographic and phenotypic data including pregnancy history and medical history can be found on FaceBase (Record ID: 56-ES6P, Accession#: FB00001368, doi: 10.25550/56-ES6P). Genotype data are available through dbGaP (accession # phs002815.v2.p1).

**POFC1 Study:** The demographic and phenotypic data are accessible through FaceBase (Record ID: 5A-FJBJ, Accession#: FB00001369, doi: 10.25550/5A-FJBJ). Genotype data are available through dbGaP (accession #: phs000774.v2.p1).

**Trios from Iowa Filipino Study genotyped via GENEVA Study**: The demographic and phenotypic data are accessible through Facebase (Record ID: 1-50DE, Accession #: FB00001040, doi: 10.25550/1-50DE)

**Trios from Iowa Filipino Study whole genome sequenced via GMKF:** The results analyzed and published here are based in part upon data generated by Gabriella Miller Kids First (GMKF) Pediatric Research Program projects phs002595.v1.p1, and were accessed from the Kids First Data Resource Portal (https://kidsfirstdrc.org/) and/or dbGaP. (www.ncbi.nlm.nih.gov/gap).

