## Supplementary material for "Gene-by-environment interactions involving maternal exposures with orofacial cleft risk in Filipinos": Figure S

### Supplemental Information

Supplemental Information includes four figures.

**Figure S1** Sample flowcharts for the discovery sample (A) and the replication samples (B).

**Figure S2** The first five principal components of ancestry (PCA) within the discovery cohort.

**Figure S3** Manhattan plots (left) and quantile-quantile (QQ) plots (right) for genome-wide GEI scan with maternal smoking in the discovery cohort.

**Figure S4** Manhattan plots (left) and quantile-quantile (QQ) plots (right) for genome-wide GEI scan with maternal vitamin use in the discovery cohort.

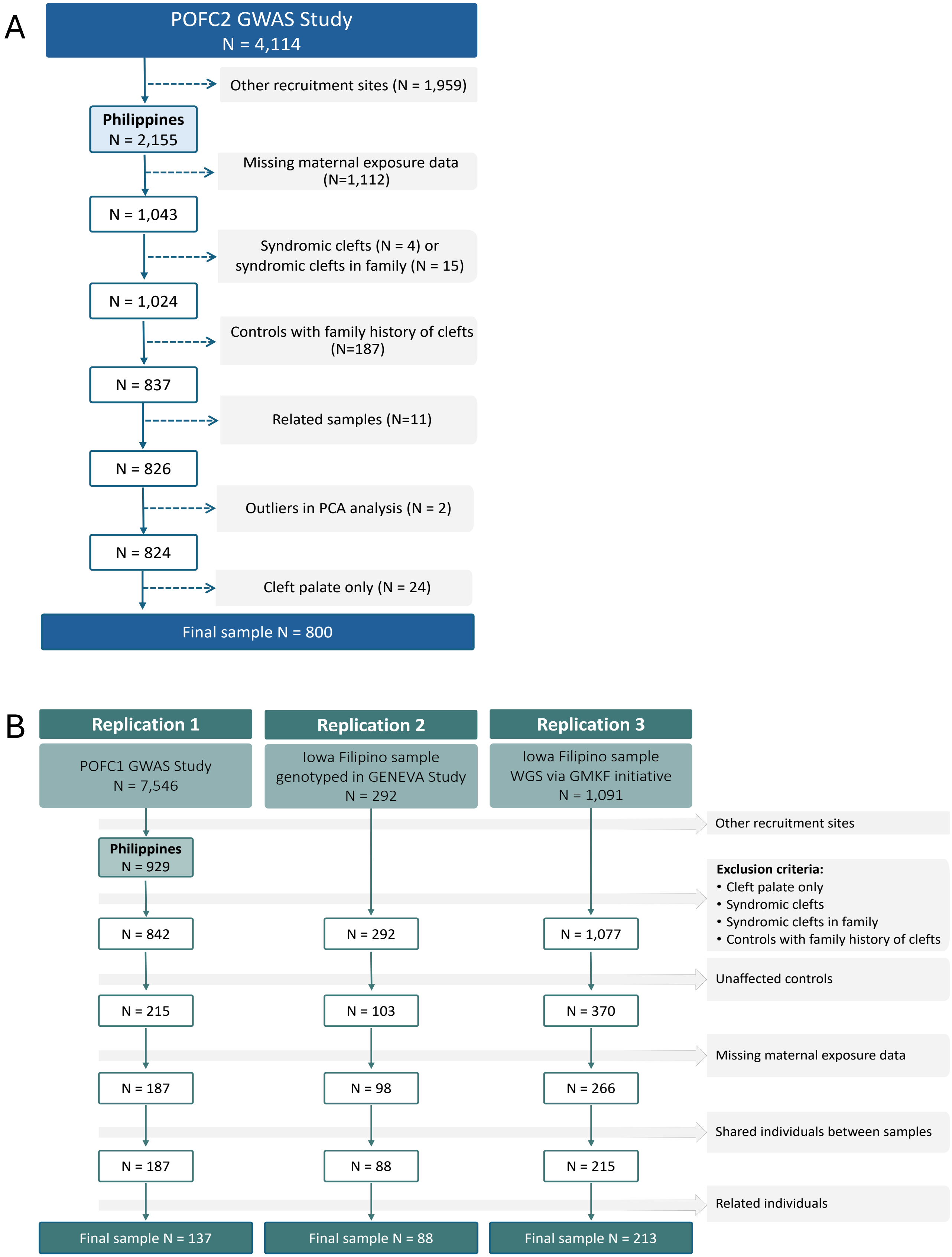


**Figure S1 Sample flowcharts for the discovery sample (A) and the replication samples (B)**

This study focused on individuals recruited in Philippines. The exclusion factors and the count of removed individuals are indicated in the light grey boxes.

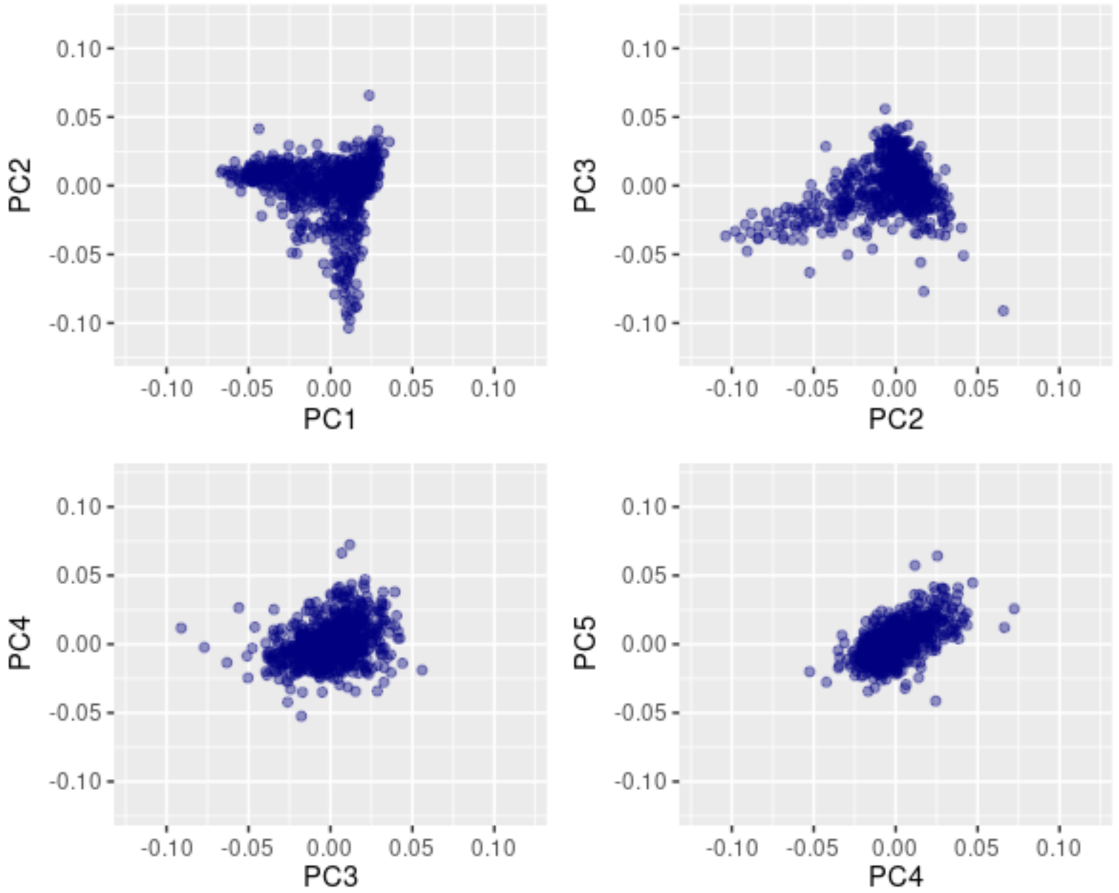


**Figure S2 The first five principal components of ancestry (PCA) within the discovery cohort.**

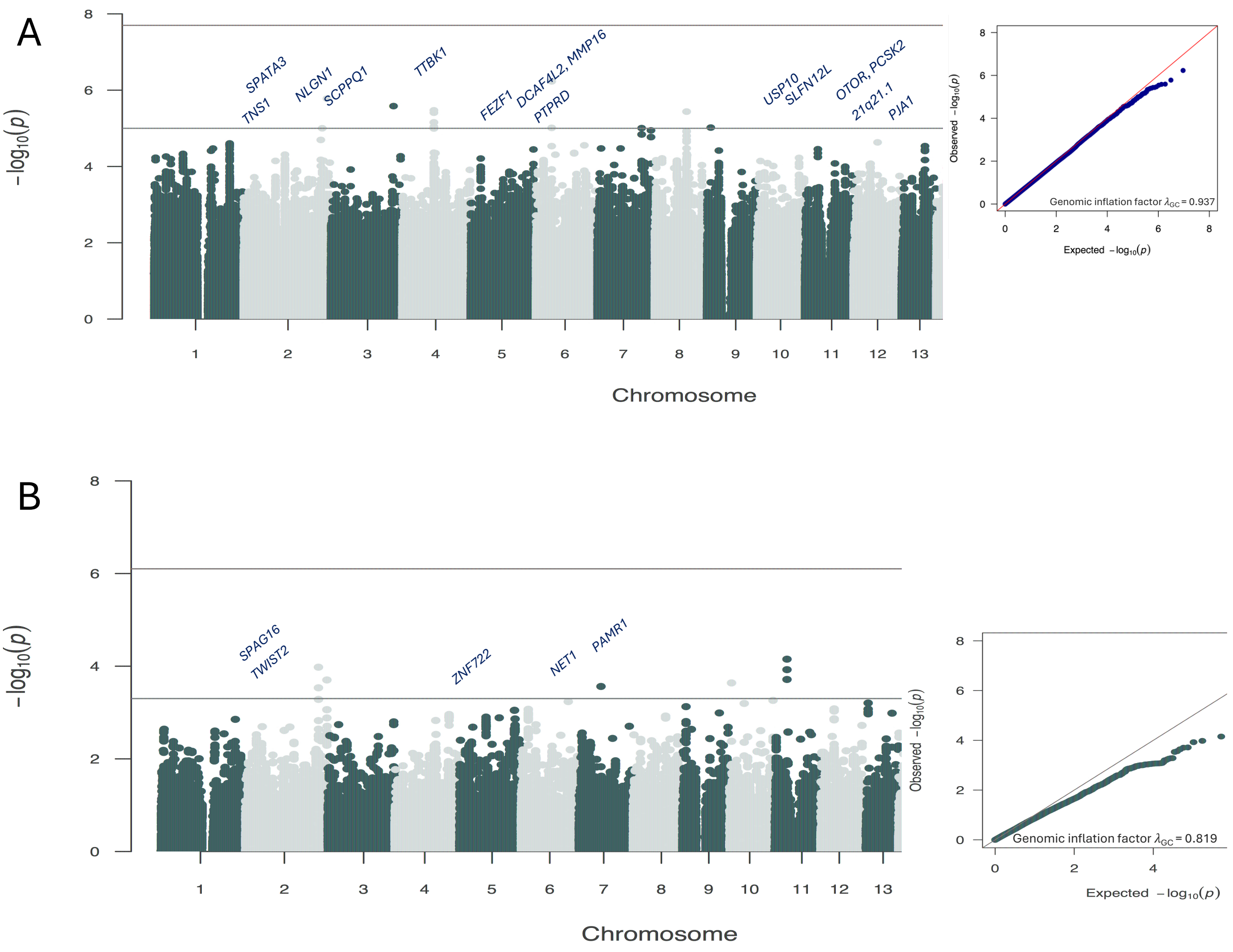


**Figure S3  Manhattan plots (left) and quantile-quantile (QQ) plots (right) for genome-wide GEI scan with maternal smoking in the discovery cohort.**

**(A)** 3df joint test with smoking and (**B**) two-step EDGE method results depicting the -$\log_{10}(p_{GxE})$ from the step 2. The red lines indicate the Bonferroni-corrected genome-wide significance thresholds for 3df test ($p_{3df}$= 5 × $10^{-8}$/3 ≈ 2 × $10^{-8}$) and two-step EDGE method ($p_{GxE}$≈ 8×$10^{-7}$), respectively. The blue lines are the suggestive thresholds $p_{3df}$= 1 × $10^{-5}$ and $p_{GxE}$ = 5 × $10^{-4}$, respectively. The lead variants are annotated with the nearby gene(s) and are mapped to the chromosomal location on the x-axis.

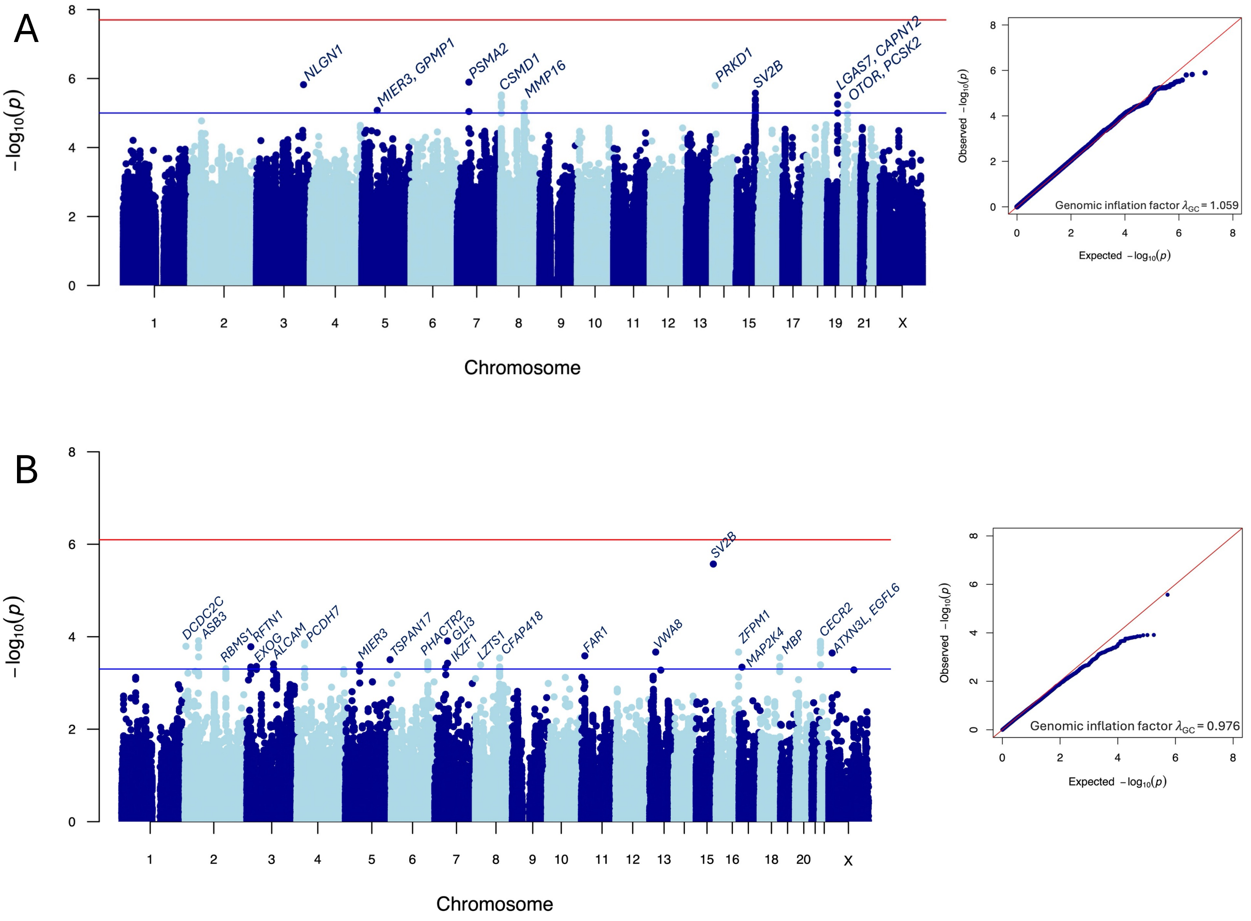


**Figure S4 Manhattan plots (left) and quantile-quantile (QQ) plots (right) for genome-wide GEI scan with maternal vitamin use in the discovery cohort.**

**(A)** 3df joint test with vitamin use and (**B**) two-step EDGE method results depicting the $-\log_{10}(p_{GxE})$ from the step 2. The red lines indicate the Bonferroni-corrected genome-wide significance thresholds for 3df test ($p_{3df} = 5 \times 10^{-8}/3 \approx 2 \times 10^{-8}$) and two-step EDGE method ($p_{GxE} \approx 8\times10^{-7}$), respectively. The blue lines are the suggestive thresholds $p_{3df} = 1 \times 10^{-5}$ and $p_{GxE} = 5 \times 10^{-4}$, respectively. The lead variants are annotated with the nearby gene(s) and are mapped to the chromosomal location on the x-axis.
